# Aortic Haemodynamics Are Independent Longitudinal Determinants of Left Ventricular Function in Repaired Tetralogy of Fallot

**DOI:** 10.64898/2026.09.11.26362746

**Authors:** Sadman Chowdhury, Panagiota Mitropoulou, Precylia Fernandes, Luca Tin Yau Cheung, Andrew M Taylor, Vivek Muthurangu, Michael A Quail

**Affiliations:** Centre for Cardiovascular Imaging, Heart and Lung Division, Great Ormond Street Hospital for Children NHS Foundation Trust, Great Ormond Street, London, UK; Research Department of Children’s Cardiovascular Disease, Institute of Cardiovascular Science, University College London, London, UK

**Keywords:** Tetralogy of Fallot, arterial compliance, systemic vascular resistance, afterload, cardiovascular magnetic resonance, congenital heart disease

## Abstract

**BACKGROUND:** Left ventricular ejection fraction (LVEF) independently predicts mortality in repaired tetralogy of Fallot (ToF), but the causes of LV dysfunction are incompletely understood. The intrinsic aortopathy of ToF increases pulsatile afterload, yet whether this contributes to LV dysfunction is unknown. We sought to determine whether aortic dimensions and flow-derived afterload act as cross-sectional markers of LV dysfunction, dynamic contributors to change in LV function over time, or both.

**METHODS:** Retrospective longitudinal study of 554 patients with repaired ToF (1,448 serial cardiovascular magnetic resonance examinations, 2003–2026). Aortic root dimensions, total arterial compliance (TAC: aortic forward flow volume/pulse pressure), and systemic vascular resistance (SVR) were derived, and a within-between decomposition separated between-patient from within-patient associations with LVEF in mixed-effects models.

**RESULTS:** In the analytic sample (1,053 observations, 521 patients), aortic sinus diameter predicted LVEF at both levels (within: β=−0.23, p<0.001; between: β=−0.20, p=0.003). Associations were reproduced at the sinotubular junction but absent at the ascending aorta, localising the effect to the root. TAC predicted LVEF within patients (β=0.90 per mL/mmHg, p=0.014) but not between patients (p=0.62), indicating that pulsatile afterload is a dynamic contributor to LV function. SVR predicted LVEF at both levels (within: β=−0.23; between: β=−0.25; both p<0.001). Blood pressure components alone, expressed as systolic and diastolic or as mean arterial and pulse pressure, did not predict LVEF and fitted the data less well. Adjustment for right ventricular ejection fraction attenuated the within-patient compliance association by 35%, suggesting a component shared with right ventricular function.

**CONCLUSIONS:** Aortic root dimensions, TAC and SVR are independent longitudinal determinants of LVEF in repaired ToF, establishing an important role for aortic haemodynamics in LV function and identifying the systemic vasculature as a potentially modifiable contributor to LV dysfunction.

**Clinical Perspective:** *What is new?:* Total arterial compliance, systemic vascular resistance and aortic root dimensions are independent longitudinal determinants of left ventricular ejection fraction in repaired tetralogy of Fallot. These associations were demonstrated within individual patients over time, indicating that aortic afterload is a dynamic, and potentially modifiable, contributor to LV function rather than a fixed cross-sectional marker.

*What are the clinical implications?:* LVEF is a powerful independent predictor of mortality in repaired tetralogy of Fallot, yet its determinants are incompletely understood. These data demonstrate a role of aortic haemodynamics in left ventricular function for the first time. The systemic vasculature should be considered alongside conventional right-sided measures when assessing patients with repaired tetralogy of Fallot, and, because pulsatile and resistive afterload are potentially modifiable, characterising the vascular phenotype may identify patients who could benefit from afterload-targeted therapy.

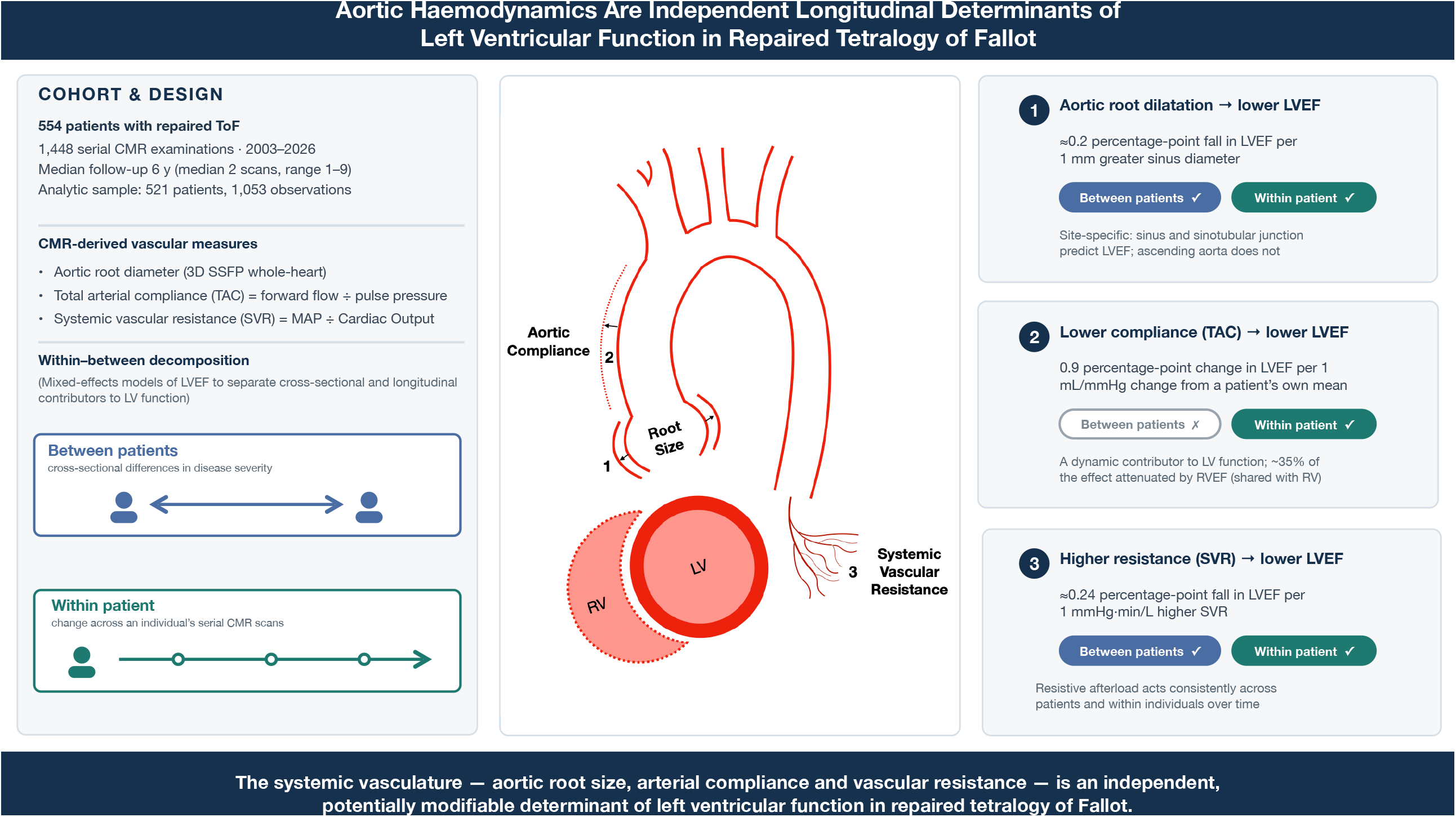

## Introduction

Tetralogy of Fallot (ToF) is the most common cyanotic congenital heart disease. Late outcomes after repair are dominated by right ventricular (RV) sequelae, yet left ventricular (LV) ejection fraction (EF) is also an independent predictor of mortality, with an effect size similar to RVEF.^1–6^

The causes of LV dysfunction are incompletely understood. A leading explanation is adverse RV-LV interaction, with measures of adverse RV remodelling associated with reduced LV systolic function.^7,8^ Aortic regurgitation, delayed repair, and prior palliative shunts have also been implicated.^8,9^

However, a substantial proportion of the variation in LV function, both between patients and within the same patient over time, remains unexplained. Patients with similar RV remodelling and comparable surgical histories can follow very different clinical courses in terms of LV dysfunction.

One source of unexplained variance that has received little attention is the systemic vasculature. The thoracic aorta in ToF is structurally and functionally abnormal.^10,11^ Medial degeneration, with elastic fibre fragmentation and smooth muscle cell loss, produces progressive root dilatation and reduced distensibility in both children and adults.^12–15^ Invasive characterisation has shown that these abnormalities translate to increased characteristic impedance, reduced total arterial compliance (TAC), elevated pulse wave velocity, and augmented wave reflection, with pulsatile load inversely correlated with cardiac index.^16^ The left ventricle therefore ejects into a stiffer, more reflective arterial system, imposing an afterload incompletely represented by conventional blood pressure measurement.

Using serial CMR in a large cohort with repaired ToF, we decomposed the associations of aortic dimensions, TAC, and systemic vascular resistance (SVR) with LVEF into between-patient and within-patient components, to determine whether these vascular measures act as cross-sectional markers of LV dysfunction, dynamic contributors to change in LV function over time, or both.

## Methods

### Study Design and Population

This was a retrospective longitudinal study of patients with repaired ToF who underwent cardiovascular magnetic resonance (CMR) imaging at Great Ormond Street Hospital for Children, between 2003 and 2026. Patients were identified from the institutional electronic health record (EHR) and included if they had undergone primary repair of ToF with at least one CMR examination.

Patients were excluded if they had ToF in association with known chromosomal abnormalities or genetic syndromes (e.g. trisomy 21, 22q11 deletion syndrome, Alagille syndrome, CHARGE syndrome). Patients were also excluded if they had ToF in combination with any of the following cardiac lesions: atrioventricular septal defect, absent pulmonary valve syndrome, pulmonary atresia, or major aortopulmonary collateral arteries.

Informed consent for the use of imaging data was obtained from all parents or guardians of the patients included in this study. The study protocol conforms to the ethical guidelines of the Declaration of Helsinki and was approved by the local committee of the UK national research ethics service (06/Q0508/124).

### Data Extraction

Structured CMR reporting data were extracted from the institutional EHR using a semi-automated tool for reading and extracting data from portable document format (PDF) reports. Extracted data were supplemented and verified by manual review of the original reports. Clinical demographic data including height, weight, body surface area, body mass index, peripheral oxygen saturation, vital status, and surgical history were obtained from the patient’s electronic record.

Surgical pulmonary valve replacement (PVR) and percutaneous pulmonary valve implantation (PPVI) were combined as a single binary exposure (pulmonary valve intervention, PVI). For each CMR study, a time-varying indicator was created denoting whether PVI had occurred before that study. A between-patient indicator of whether the patient had ever undergone PVI during the study period was also derived.

### Cardiac Magnetic Resonance Imaging

CMR was performed using 1.5-T MR scanners (Siemens Healthcare, Erlangen, Germany) according to institutional protocols. During the study period, the MRI scanner hardware was upgraded from “Avanto” to “Avanto fit”.

#### Ventricular Volumes and Function

Biventricular volumes and function were assessed using cine imaging in the short-axis orientation covering both ventricles. Left ventricular end-diastolic volume, end-systolic volume, stroke volume, and ejection fraction (LVEF) were calculated from manual segmentation of short-axis cine images at end-diastole and end-systole using Simpson’s rule. Right ventricular (RV) volumes and ejection fraction (RVEF) were derived using the same method. Throughout the study period LV and RV trabeculations were excluded from the blood pool.

#### Aortic and Pulmonary Artery Flow

Phase-contrast flow data were acquired in the ascending aorta (positioned just above the sinotubular junction) and main pulmonary artery using velocity-encoded gradient-echo sequences. Forward flow, backward flow, net flow, and regurgitant fraction were calculated for both the aortic and pulmonary positions from phase-contrast images using a semi-automatic vessel edge-detection algorithm.

Residual pulmonary stenosis was graded as none, mild, moderate or severe based on velocity on phase contrast imaging in combination with anatomic narrowing on 3D and cine imaging.

#### Aortic Root Dimensions

Aortic root dimensions were measured from three-dimensional SSFP whole-heart imaging, which has been used at our institution throughout the study period. Three aortic sinus diameters, two orthogonal sinotubular junction (STJ) diameters, and two orthogonal ascending aortic diameters were measured. The mean sinus diameter was calculated as the average of three measurements. The mean STJ and ascending aortic diameters were each calculated as the average of two measurements.

#### Blood Pressure

Blood pressure measurement was not routinely recorded for all patients during CMR assessment prior to 2009. In patients in whom it was recorded, brachial systolic (SBP) and diastolic (DBP) blood pressure were measured by automated oscillometric sphygmomanometry immediately before the CMR examination. Pulse pressure was calculated as SBP minus DBP. Mean arterial pressure (MAP) was obtained from the oscillometric device where available or calculated as DBP + (SBP − DBP)/3 when only systolic and diastolic pressures were recorded.

Office readings were classified as normal, elevated, or hypertensive range using age-appropriate criteria (Supplemental Methods); as blood pressure was a single office measurement at each visit, these categories represent elevated office readings rather than diagnosed hypertension.

### Derived Measurements

#### Total Arterial Compliance

TAC was calculated as aortic forward flow volume, SV (phase-contrast derived, ml) divided by pulse pressure (SV/PP), unit: ml/mmHg.

#### Systemic Vascular Resistance

SVR was calculated as mean arterial pressure divided by aortic flow (phase-contrast derived flow, L/min), unit: mmHg·min/L.

### Statistical Analysis

All analyses were performed using Stata 19.5 (StataCorp, College Station, TX). Continuous data are expressed as mean (95% confidence interval) or median (interquartile range) as appropriate. A two-sided significance level of 0.05 was used throughout.

#### Mixed-Effects Regression

The association between vascular indices and LVEF was assessed using linear mixed-effects regression with maximum likelihood estimation,^17^ with a patient-level random intercept to account for repeated measurements.

To distinguish cross-sectional from longitudinal associations, a within-between (hybrid) decomposition was applied to all time-varying covariates (TAC, SVR, mean sinus diameter, RVEF, pulmonary regurgitant fraction, aortic regurgitant fraction, and pulmonary stenosis severity). Each of these covariates was separated into a between-patient component (the patient’s mean across observations) and a within-patient component (the deviation from that mean), entered simultaneously. Sex, body surface area, and age at study were entered as time-invariant fixed effects. Differences between within-patient and between-patient coefficients were assessed by post-estimation Wald tests, a significant difference indicating that a single pooled effect would inadequately summarise the association.

The relative contribution of between- and within-patient variation for each time-varying covariate was summarised by the intraclass correlation coefficient (ICC) from a random-intercept model, to confirm that sufficient within-patient variation was available to estimate longitudinal associations. Model performance was summarised by marginal and conditional R^2^ (variance explained by fixed effects, and by fixed plus random effects, respectively) and by the proportional reduction in between- and within-patient variance relative to a random-intercept-only model. Two further model comparisons were performed. The primary model was refitted without RVEF, to estimate the total effect of TAC on LVEF including any component shared with RV function. It was also refitted with TAC and SVR replaced by decomposed brachial pressure components, expressed as systolic and diastolic blood pressure (SBP, DBP) or as mean arterial and pulse pressure (MAP, PP), to test whether the derived measures captured vascular load better than blood pressure alone. These substitution models are not nested within the primary model, so fit was compared using the Akaike and Bayesian information criteria (AIC, BIC) on the identical estimation sample, with lower values indicating better fit.

Robustness was assessed in five prespecified sensitivity analyses (adjustment for study era and for pulmonary valve intervention status, substitution of sinotubular junction and ascending aortic diameters, and calibration for a sinus measurement-convention drift); assessment of temporal trends and handling of missing data are described in the Supplemental Methods.

## Results

### Study Population

554 patients with repaired tetralogy of Fallot contributed 1,448 CMR examinations performed between 2003 and 2026, Table 1). Patients underwent a median of 2 examinations (interquartile range 1–4, range 1–9) over a median follow-up of 6 years (interquartile range 0–12). 392 patients (71%) had two or more examinations. Median age at first study was 16 years (interquartile range 11–32); 312 (56%) were male. Mean LV end-diastolic volume index at first CMR was 73 ± 15 mL/m^2^ and mean LVEF was 62 ± 7%. Mean RV end-diastolic volume index was 112 ± 34 mL/m^2^ and mean RVEF was 57 ± 9%, with a mean pulmonary regurgitant fraction of 28 ± 15%. 146 patients (26%) underwent pulmonary valve intervention during the study period, of whom 79 (54%) had CMR examinations both before and after the procedure. 18 patients (3%) died during follow-up.

**Table 1.**
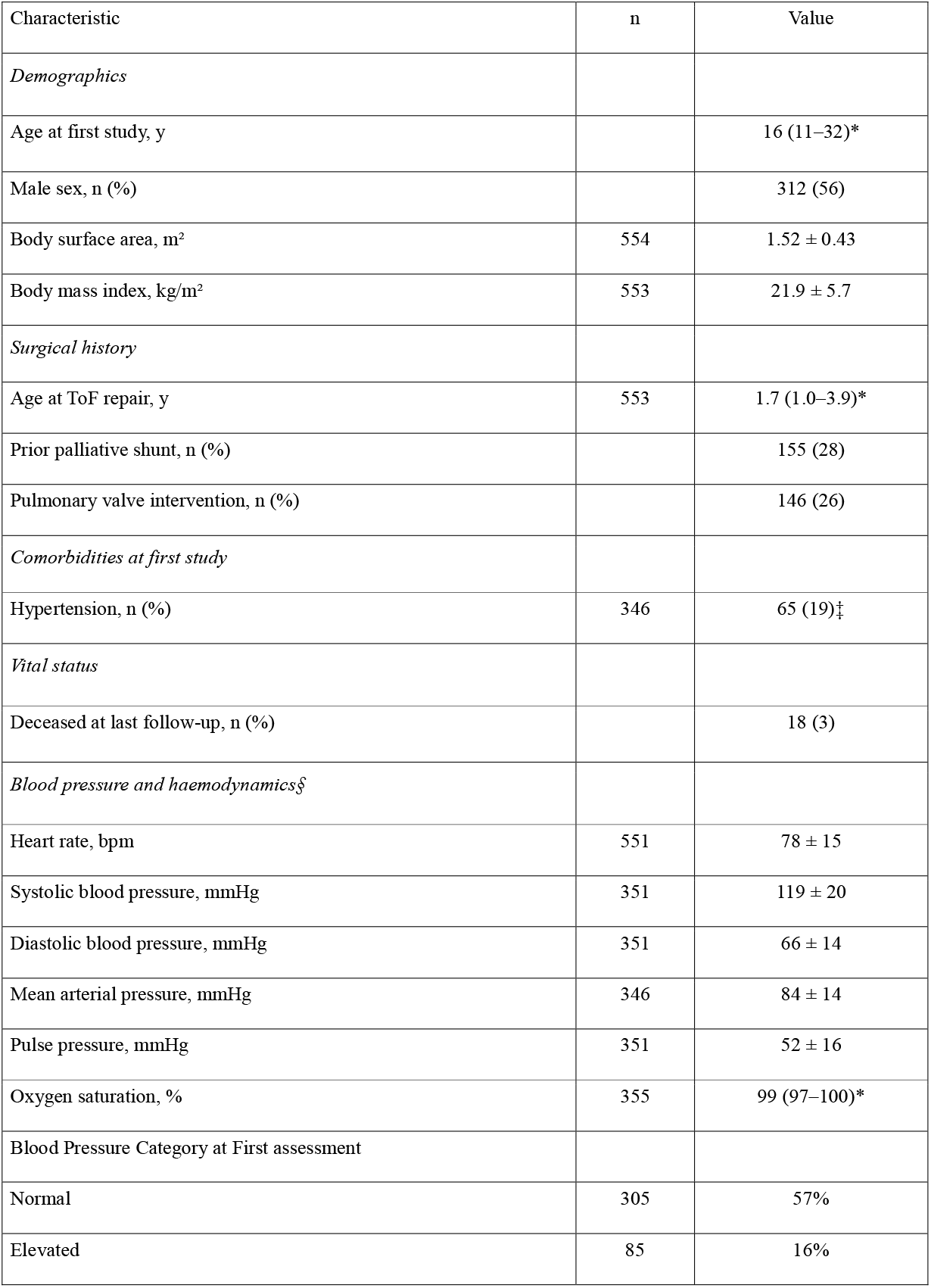
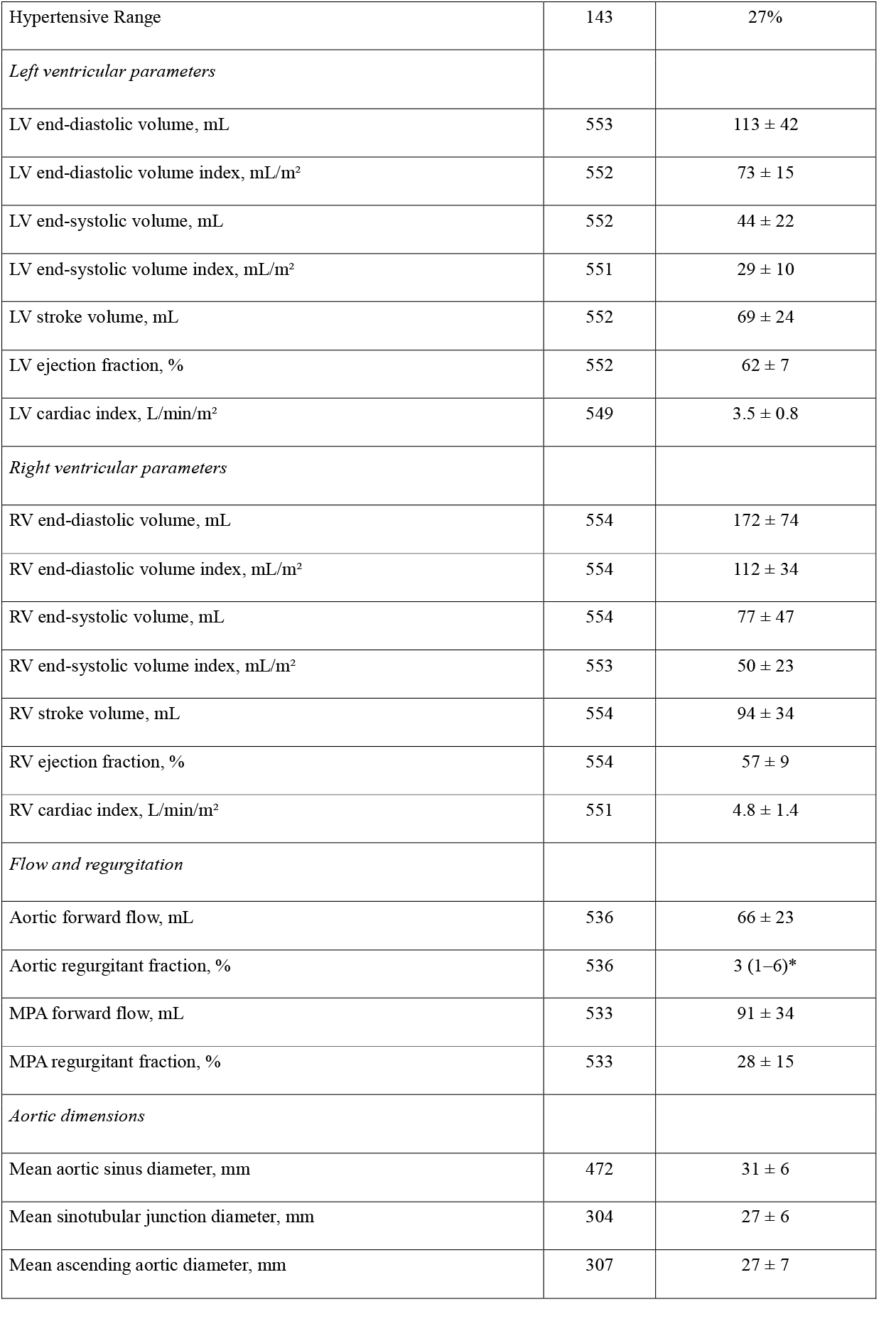
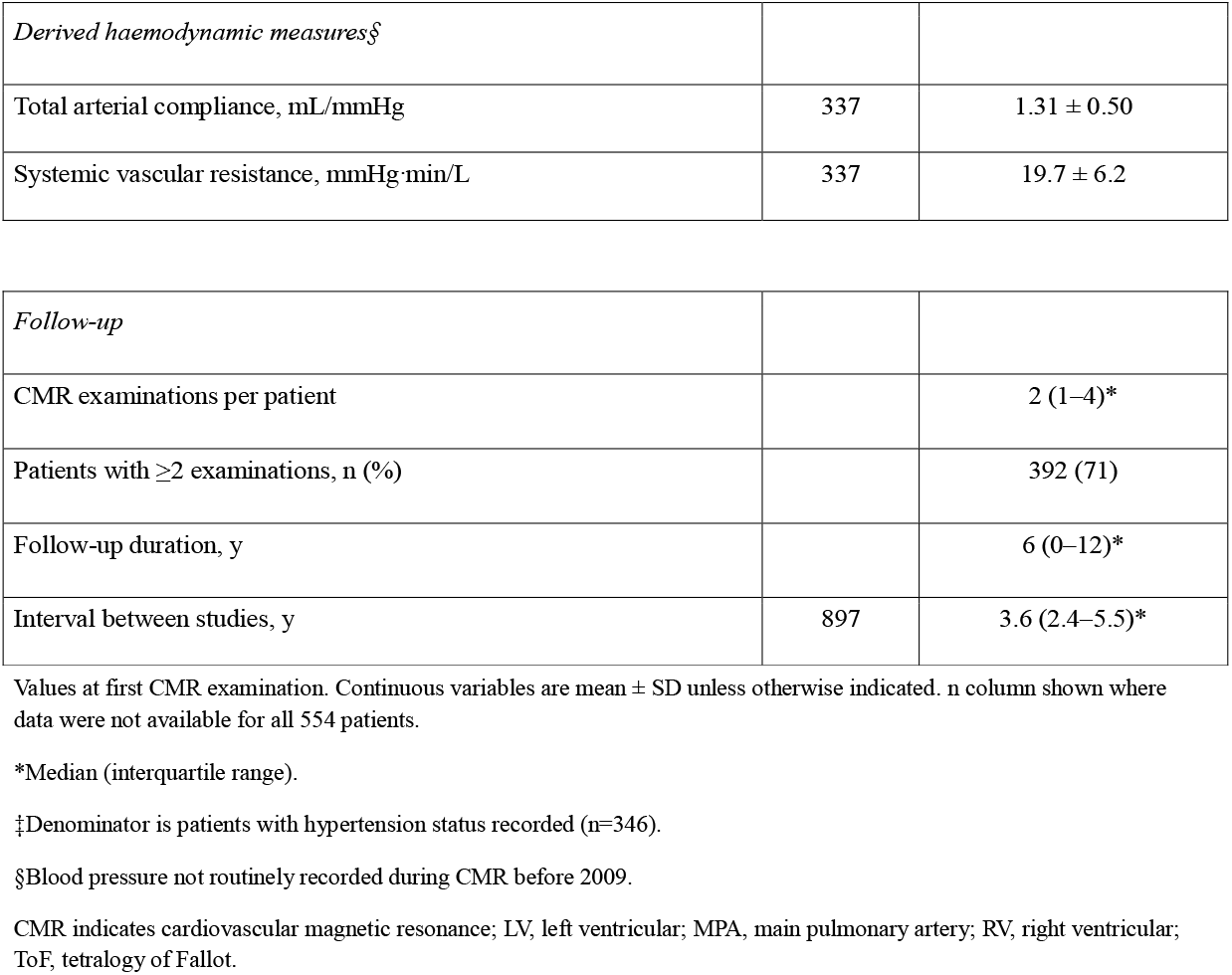
Baseline Characteristics of the Study Population.

Prior to 2009, BP was not routinely recorded during CMR assessment, and consequently TAC and SVR could not be derived for a significant proportion of earlier studies. The analytic sample for the primary model therefore comprised 1,053 observations from 521 patients. Patients with and without compliance data did not differ in LVEF, age, or sex (p=0.8).

Elevated blood pressure was common. At first assessment, 143 of 533 patients (27%) had blood pressure in the hypertensive range, although isolated elevated readings may partly reflect the measurement setting. More robustly, blood pressure remained in the hypertensive range at more than half of visits in 190 patients (36%), suggesting sustained elevation in a substantial minority.

The three vascular exposures showed distinct trajectories across age (Figure 1): sinus diameter rose through childhood before plateauing, TAC peaked in early adulthood and declined thereafter, and SVR fell to an adolescent nadir before rising. Together with the ICCs (Supplemental Table 1), these confirm substantial within-patient variation over time, the prerequisite for estimating longitudinal associations.

**Figure 1.**
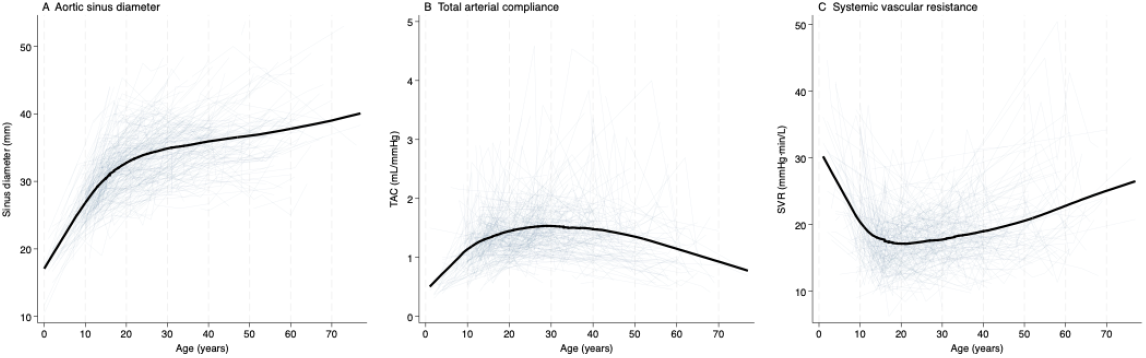
Trajectories of vascular exposures across age in repaired tetralogy of Fallot. Faint lines show individual patient trajectories for (A) aortic sinus diameter, (B) total arterial compliance, and (C) systemic vascular resistance, plotted against age at study. The bold line shows the population trend (locally weighted regression). Aortic sinus diameter rises steeply through childhood and adolescence before plateauing in adulthood. Total arterial compliance increases through childhood, peaks in early adulthood, and declines thereafter. Systemic vascular resistance falls during childhood, reaches a nadir in adolescence, and rises progressively through adult life. The visual spread illustrates that the relative contribution of between-patient and within-patient variation differs across the three measures.

### Associations With Left Ventricular Ejection Fraction

The results of the regression model are presented in Table 2 and Figure 2. Both the pulsatile and resistive components of vascular afterload were independently associated with LVEF, but with distinct patterns of association.

**Table 2.** Within-Between Decomposition of Associations With Left Ventricular Ejection Fraction.

| Covariate | Between-patient |  | Within-patient |  | Wald P |
| --- | --- | --- | --- | --- | --- |
| | $\beta$ (95% CI) | P | $\beta$ (95% CI) | P | |
| Compliance | -0.30 (-1.51, 0.90) | 0.62 | 0.90 (0.18, 1.62) | 0.014 | 0.091 |
| SVR | -0.25 (-0.37, -0.14) | <0.001 | -0.23 (-0.32, -0.15) | <0.001 | 0.80 |
| Sinus diameter | -0.20 (-0.33, -0.07) | 0.003 | -0.23 (-0.35, -0.10) | <0.001 | 0.70 |
| RVEF | 0.43 (0.37, 0.50) | <0.001 | 0.38 (0.31, 0.45) | <0.001 | 0.31 |
| Pulmonary RF | 0.006 (-0.03, 0.04) | 0.72 | -0.04 (-0.07, 0.001) | 0.059 | 0.10 |
| Aortic RF | -0.09 (-0.19, 0.02) | 0.11 | -0.05 (-0.14, 0.05) | 0.35 | 0.60 |
| Sex (male) | 0.05 (-0.93, 1.04) | 0.92 |  |  |  |
| BSA | -0.23 (-1.90, 1.43) | 0.78 |  |  |  |
| Age at study | 0.05 (0.01, 0.09) | 0.022 |  |  |  |

1,053 observations from 521 patients. $\beta$ coefficients represent the change in LVEF (%) per unit increase in each covariate. Between-patient coefficients represent cross-sectional associations; within-patient coefficients represent longitudinal associations within individuals. Wald P tests equality of between-patient and within-patient coefficients; a significant result indicates the pooled coefficient is an inadequate summary. BSA indicates body surface area; LVEF, left ventricular ejection fraction; MPA RF, main pulmonary artery regurgitant fraction; RF, regurgitant fraction; RVEF, right ventricular ejection fraction; SVR, systemic vascular resistance.

**Figure 2.**
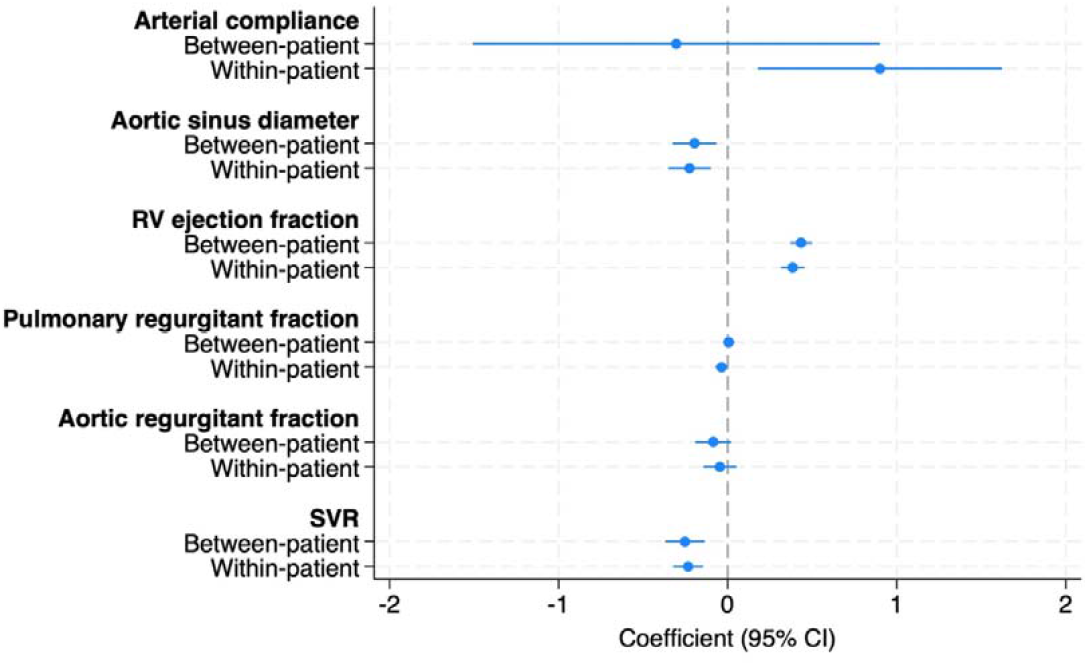
Between-patient and within-patient coefficients from the primary mixed-effects model. Each covariate is decomposed into between-patient (cross-sectional) and within-patient (longitudinal) components. Points represent β coefficients with 95% confidence intervals. The dashed vertical line indicates null effect. Total arterial compliance shows a significant within-patient effect with a null between-patient effect, indicating that pulsatile afterload is a dynamic contributor to LV function. Systemic vascular resistance shows equivalent effects at both levels. 1,053 observations from 521 patients.

Mean aortic sinus diameter was associated with lower LVEF at both levels: each 1 mm greater diameter corresponded to approximately 0.2 percentage points lower LVEF between patients and 0.23 points within patients (both p≤0.003; Wald p=0.70; full coefficients in Table 2). Anatomical specificity was assessed by substituting the sinus measurement with adjacent aortic segments. The sinotubular junction showed comparable associations: each 1 mm greater diameter corresponded to approximately 0.18 percentage points lower LVEF between patients (p=0.009) and 0.26 points within patients (p=0.016). In contrast, ascending aortic diameter was not associated with LVEF at either level (p=0.84 and p=0.10 respectively). This localises the association to the aortic root.

TAC was associated with LVEF within patients but not between patients. Specifically, each 1 mL/mmHg rise in compliance from a patient’s own average corresponded to a significant (p=0.014) 0.9-percentage-point increase in LVEF, whereas the between-patient coefficient was non-significant (p=0.62). Full coefficients and confidence intervals are given in Table 2. The within-versus between-patient Wald test approached significance (p=0.09); together with the opposing directions of the two coefficients, this indicates an association driven by within-patient change rather than cross-sectional differences.

In contrast, SVR was associated with LVEF at both levels with each 1 mmHg·min/L higher SVR corresponding to a fall in LVEF of about 0.25 percentage points between patients and 0.23 within patients (both p<0.001). The two coefficients were statistically equivalent (Wald p=0.80; Table 2), indicating that the resistive component of afterload operates consistently whether comparing patients or tracking individuals over time.

Substituting blood pressure components for the derived haemodynamic measures, no component (SBP, DBP, MAP, PP) was independently associated with LVEF either within or between patients (p≥0.13). On the same 1,053 observations, the model using the derived measures of TAC and SVR fitted substantially better than either blood-pressure model, which fitted near-identically to each other (AIC 6688 versus 6752; BIC 6767 versus 6831), indicating that pressure scaled by aortic flow captures vascular load more completely than cuff pressure alone.

The within-patient marginal effects of aortic sinus diameter, TAC, and systemic vascular resistance on predicted LVEF are illustrated in Figure 3.

**Figure 3.**
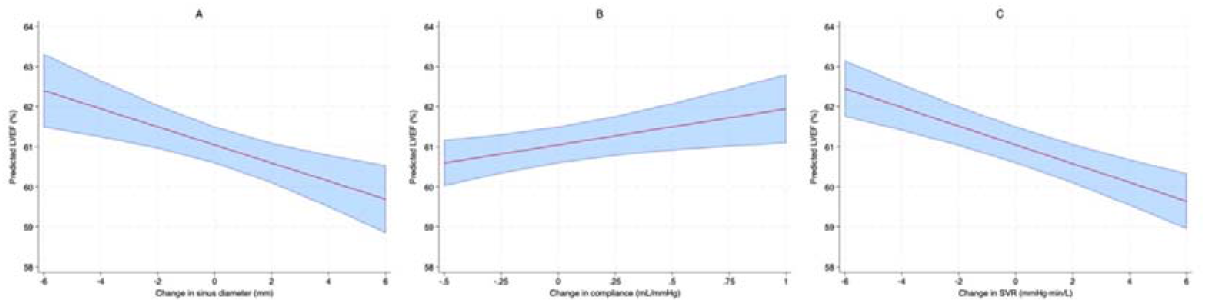
Predicted LVEF as a function of within-patient change in (A) aortic sinus diameter, (B) total arterial compliance, and (C) systemic vascular resistance. Each panel shows the marginal prediction from the primary model, with all other within-patient covariates held at their mean (zero deviation). Shaded areas represent 95% confidence intervals. The x-axis represents deviation from each patient’s own mean, illustrating the predicted LVEF change when an individual patient’s covariate value changes over time.

Aortic regurgitant fraction was not independently associated with LVEF at either level (p≥0.11; Table 2). Pulmonary regurgitant fraction showed a within-patient association that approached significance (p=0.059) with a null between-patient effect (p=0.72), and the grading of residual pulmonary stenosis severity was non-significant at either level (p≥0.47). Age at study was positively associated with LVEF (β=0.05, p=0.02), whereas sex and body surface area were not significant. Age at primary repair and prior palliative shunt were tested as additional covariates but were not significant (p=0.21 and p=0.92 respectively) and did not alter the primary findings.

RVEF was strongly associated with LVEF at both levels, with comparable between- and within-patient coefficients (β=0.43 and 0.38; both p<0.001; Wald p=0.31; Table 2).

#### Variance Explained

The covariates accounted for 42% of between-patient differences in LVEF and 26% of within-patient visit-to-visit change (marginal R^2^=0.35; conditional R^2^=0.72; full variance decomposition in Supplemental Results). Substantial residual between-patient variance indicates that unmeasured stable patient characteristics also contribute.

#### Effect of RVEF Adjustment on Vascular Afterload

In the primary model, the within-patient TAC coefficient was 0.90. Removing RVEF increased it to 1.39 (p=0.001); equivalently, adjusting for RVEF attenuated the coefficient by 35% (from 1.39 to 0.90). This indicates that roughly one-third of the total within-patient compliance effect on LVEF is shared with pathways involving RV function, either through ventricular interdependence or through a common mechanism affecting both ventricles. By contrast, the within-patient coefficients for SVR (−0.24 versus −0.24) and aortic sinus diameter (−0.23 versus −0.24) were unchanged by the removal of RVEF, indicating that the resistive afterload and aortic root effects on LVEF are independent of RV function.

#### Sensitivity Analyses

All primary findings were unchanged in sensitivity analyses adjusting for study era or pulmonary valve intervention status (Supplemental Results). A gradual drift in sinus measurement convention was identified (sinus-to-STJ ratio +0.006 per year, p<0.001); after calibration, the within-patient sinus coefficient was modestly strengthened (β=−0.29, p<0.001), with all other coefficients unchanged.

## Discussion

In this longitudinal study of patients with repaired tetralogy of Fallot, we used a within–between decomposition to separate cross-sectional from longitudinal associations between aortic geometry, haemodynamics and left ventricular function. The principal findings were that (1) aortic root dimensions independently predicted LVEF at both the between-patient and within-patient levels, with anatomical site specificity (a significant association at the root but a null association at the ascending aorta); (2) TAC predicted LVEF within patients but not between patients, indicating that pulsatile afterload is a dynamic contributor to LV function; (3) systemic vascular resistance predicted LVEF consistently at both levels; and (4) the within-patient compliance effect was partly attenuated by adjustment for RVEF, whereas the root-dimension and SVR effects were independent of RV function, suggesting that the compliance association may be partly shared with the right ventricle. To our knowledge, this represents the largest serial CMR cohort in repaired ToF reported to date (1,448 examinations in 554 patients, 392 with serial imaging, over 20 years).

Aortic root dilatation is a characteristic feature of ToF.^10^ Whilst a dilated aortic root can contribute to the development of aortic regurgitation, it has hitherto been considered a relatively benign component of the condition; particularly because of a low risk of acute aortic events including during higher risk periods such as pregnancy.^18^

Histological studies of the aorta in ToF have shown medial degeneration, elastic fibre fragmentation, and smooth muscle cell loss. These structural changes translate to important alteration of arterial wall biomechanics, manifesting as increased aortic stiffness, larger aortic dimensions, and higher impedance. Collectively, these findings support the concept of a diffuse vasculopathy in ToF that could contribute to increased pulsatile load and adverse ventriculo–arterial coupling.^16^

Our study has demonstrated that aortic root dimensions (sinus or STJ) are independently associated with LVEF at both the between-patient and within-patient levels with equal magnitude. These data indicate that root dilatation is both a severity marker and a longitudinal predictor of LVEF.

The relationship between root size and LV function is complex. Localised aortic root dilatation can produce an impedance mismatch with a smaller-calibre ascending aorta, generating wave reflections that increase LV afterload, ^19,20^ while wall stiffening and increased pulse wave velocity oppose the fall in characteristic impedance expected from a larger lumen.^16,20^ Abnormal root biomechanics, together with reduced TAC and altered wave reflections, may also adversely influence coronary perfusion.^21,22^

### Pulsatile Versus Resistive Afterload

In addition to root geometry, we also derived the major components of pulsatile and resistive afterload using phase-contrast flow MRI and contemporaneous blood pressure: TAC and SVR. These two components showed distinct patterns of association with LVEF. TAC was associated with LVEF only within patients: when an individual patient’s compliance changed over time, their LVEF changed correspondingly. In contrast, SVR was associated with LVEF at both levels, with equivalent between-patient and within-patient coefficients. The resistive component of afterload therefore operates consistently whether comparing patients cross-sectionally or tracking individuals longitudinally. The absence of a between-patient association may reflect residual confounding: between-patient differences in TAC are strongly determined by body size, age, and aortic geometry, and the opposite sign of the between-patient coefficient is consistent with incomplete adjustment for these stable characteristics. Within-patient change removes such confounding by design, so the within-patient estimate more directly reflects the effect of altered pulsatile load.

Brachial blood pressure components, whether expressed as SBP and DBP or as MAP and PP, were not independently associated with LVEF when substituted for the derived measures, and gave a poorer overall fit by both information criteria. This does not mean pressure is irrelevant: TAC and SVR are themselves derived in part from pulse pressure and mean arterial pressure. Rather, it indicates that pressure alone is insufficient, and that the association with LV function emerges only when pressure is scaled by aortic flow to characterise the pulsatile (compliance) and resistive (resistance) components of afterload. Specific assessment of these measures, rather than cuff pressure alone, therefore appears necessary to capture the vascular load imposed on the left ventricle in this population.

### Relationship to Right Ventricular Function

RVEF was the strongest predictor of LVEF in this study, with consistent effects at both levels (between: 0.43; within: 0.38). This is conventionally attributed to ventricular interdependence, the mechanical coupling between the ventricles through the interventricular septum, the shared pericardium, and common myocardial fibres. Consistent with this, we have previously shown that pulmonary valve replacement increased LV stroke volume and output only in patients with severe RV dilatation and reduced cardiac output, an effect attributable to restored LV filling, whereas LV indices were unchanged in those with moderate dilatation;^23^ a recent serial CMR analysis similarly found that left-sided measures of structure and function were associated with rates of RV remodelling.^24^

Thirty-five percent of the within-patient compliance effect on LVEF was attenuated by adjustment for RVEF, whereas the SVR and root-dimension effects were unchanged. If compliance affected the LV exclusively through direct afterload, adjustment for RVEF should not alter its coefficient: the RV ejects into the pulmonary circulation, not the aorta. The attenuation instead suggests that compliance changes are accompanied by parallel changes in both ventricular ejection fractions. This evidence is indirect and hypothesis-generating: candidate explanations, including a common myocardial or coronary process affecting both ventricles or shared developmental severity, cannot be distinguished in the present data.

An alternative explanation for the shared compliance association is confounding by a common developmental factor. The severity of the ToF aortopathy may track with the overall severity of the cardiac malformation; patients with more severe developmental abnormalities may have both worse aortic wall properties and worse myocardial resilience, without a causal link between the two. Whilst the within-patient design mitigates cross-sectional confounding, it cannot exclude the possibility that compliance and ventricular function deteriorate in parallel because of a shared underlying trajectory.

Taken together with the structural findings, these results suggest that the aortopathy of ToF may affect LV function through more than one pathway: a direct structural effect of root dilatation on the local haemodynamic environment, and an effect of reduced compliance whose partial attenuation by RVEF raises the possibility of a process shared with the right ventricle. Prospective studies are required to define these mechanisms.

### Covariates not associated with LVEF

Notably, the lesions that dominate surveillance in repaired ToF were not independent determinants of LVEF. Pulmonary regurgitation and residual pulmonary stenosis showed no independent association, and aortic regurgitation, despite volume-loading the LV, was likewise not independently associated once vascular load was accounted for. LV function in this population appears to be governed more by the systemic vascular load than by the right-sided and valvar lesions that currently anchor follow-up.

### Clinical Implications

These findings suggest that the systemic vasculature, and specifically the aortic root, should be considered alongside conventional cardiac measures when assessing patients with repaired ToF. Current surveillance focuses on RV volumes, pulmonary regurgitation, and arrhythmia burden; the aortic root and TAC receive relatively little clinical attention despite the well-documented aortopathy in this population.

Pulsatile and resistive afterload are potentially modifiable pharmacologically and through aerobic exercise.^25^ Two randomised trials of renin-angiotensin system inhibition in repaired ToF (APPROPRIATE, ramipril; REDEFINE, losartan) were neutral for their primary endpoints, but each identified subgroups with evidence of LV benefit, and neither assessed TAC or vascular function as part of the treatment response.^26,27^

Our finding that total arterial compliance is an independent and dynamic determinant of LV function, with a within-patient effect that was partly attenuated by RV function, suggests that characterising the vascular phenotype may be important both for understanding variable treatment responses in these trials and for identifying patients most likely to benefit from vascular-targeted therapy. Prospective studies incorporating direct assessment of arterial haemodynamics and vascular function are needed before treatment can be appropriately targeted in this population.

### Limitations

Several limitations should be considered. This was a retrospective analysis of routinely acquired clinical data, subject to the limitations inherent in electronic health record studies, including variability in measurement protocols, multiple observers, and incomplete data.

Blood pressure was not routinely recorded before 2009, limiting the analytic sample for compliance and SVR analyses to 521 of 554 patients; however, patients with and without compliance data did not differ in LVEF, age, or sex.

TAC was estimated as aortic forward flow volume divided by pulse pressure, a simplified measure that does not capture the frequency dependence of arterial impedance; windkessel modelling would provide a more physiologically grounded estimate. We also did not assess pulse wave velocity or wave reflections, further important components of pulsatile load that, given the root morphology in this population, may contribute independently to LV loading.

Aortic sinus dimensions were measured by multiple observers over the 20-year study period. A gradual shift in measurement convention was identified through examination of the sinus-to-sinotubular junction ratio over time and was addressed through statistical calibration and sensitivity analyses substituting the sinotubular junction, which demonstrated consistent associations with LVEF.

The within-between decomposition estimates average within-patient effects across the population and cannot identify individual patients in whom these associations are strongest or weakest. Whilst the within-patient design substantially reduces confounding by stable patient characteristics compared with cross-sectional analyses, it cannot establish causation. Unmeasured time-varying confounders, including changes in medication or physical activity may contribute to the observed associations. Antihypertensive medication data were not systematically available.

Finally, whilst routine CMR provides accurate quantification of ventricular volumes, flow, and regurgitation, we did not systematically quantify myocardial fibrosis, RV/LV mass or measure diastolic function, all of which may contribute to the residual between-patient variance in LVEF observed in this study.

## Conclusions

In this large longitudinal study of patients with repaired tetralogy of Fallot, aortic root dimensions, TAC and SVR were independent longitudinal determinants of left ventricular function. Root dilatation predicted LVEF both cross-sectionally and longitudinally, with anatomical specificity at the aortic root, and reduced compliance was a dynamic within-patient predictor of LVEF that was partly attenuated by RV function. These findings identify the systemic vasculature as an important and potentially modifiable contributor to left ventricular function in repaired tetralogy of Fallot.

## Data Availability

All data produced in the present study are available upon reasonable request to the authors

## Abbreviations

CMR: cardiovascular magnetic resonance
LVEF: left ventricular ejection fraction
MAP: mean arterial pressure
PP: pulse pressure
PVI: pulmonary valve intervention
RVEF: right ventricular ejection fraction
STJ: sinotubular junction
SVR: systemic vascular resistance
TAC: total arterial compliance
ToF: tetralogy of Fallot

## Declaration of AI-Assisted Technologies

During the preparation of this manuscript, the authors used a large language model (Claude, Anthropic) to assist with checking stylistic consistency with journal author requirements; identifying spelling and grammatical errors; verifying reference formatting; cross-checking reported values in the text and tables against the original Stata output; generating formatted tables from that output; and shortening author-written sections to comply with word limits. All AI-assisted output, including any shortened text and formatted tables, was reviewed, verified against the original statistical output, and edited by the authors. All conceptualisation, data collection, data analysis, interpretation of findings, and clinical and mechanistic conclusions were undertaken by the authors, who take full responsibility for the content of this article.

