## Supplement for "Aortic Haemodynamics Are Independent Longitudinal Determinants of Left Ventricular Function in Repaired Tetralogy of Fallot"

**Supplemental Material**

**Supplemental Methods**

*Blood Pressure Classification*

Blood pressure was classified using age-appropriate criteria. In children under 13, systolic and diastolic pressures were converted to age-, sex-, and height-specific percentiles using the regression coefficients of the Fourth Report (NHLBI 2005), with height-for-age z-scores from the CDC 2000 reference; values at or above the 95th percentile were classified as hypertensive range and the 90th to 95th percentile as elevated. Adolescents aged 13 to 17 were classified using static AAP 2017 thresholds and adults using 140/90 mmHg.^1,2^ As blood pressure was a single office measurement at each visit, these categories represent elevated office readings rather than diagnosed hypertension.

*Sensitivity Analyses*

The robustness of findings was assessed through the following sensitivity analyses: (1) adjustment for four study eras (pre-2010, 2010–2014, 2015–2019, 2020 onwards); (2) adjustment for pulmonary valve intervention status; (3) substitution of the mean sinus diameter with sinotubular junction diameter, measured at unambiguous anatomical landmarks not subject to measurement convention variability; (4) substitution with ascending aortic diameter, to assess whether associations were specific to the aortic root; and (5) correction of sinus dimensions for a measurement convention drift identified over the study period (see below).

*Assessment of Temporal Bias*

To evaluate whether calendar trends in measured variables could confound the primary findings, body surface area-adjusted temporal trends were examined for all key covariates. A gradual shift in sinus measurement convention was identified by examining the ratio of sinus diameter to sinotubular junction diameter over calendar time. As a sensitivity analysis, sinus dimensions were calibrated using a linear regression of the sinus-to-STJ ratio on study year, with all measurements standardised to the ratio expected at the median study year (2016).

Missingness

Patients with and without available compliance data were compared on baseline LVEF, RVEF, age, sex, and pulmonary valve intervention status. The mixed-effects model uses all available observations for each patient; patients were excluded from a given analysis only if the relevant covariates were missing at all time points.

**Supplemental Results**

*Variance Explained*

Patients underwent repeated imaging, so variation in LVEF has two components: stable differences between patients, and visit-to-visit change within patients. Between patients, the random-intercept variance fell from 32.1 in the null model to 18.7 in the full model, meaning the covariates accounted for 42% of the systematic differences in LVEF between individuals. Within patients, the residual variance fell from 18.8 to 13.9, meaning 26% of the visit-to-visit change in an individual's LVEF was accounted for by concurrent changes in the measured covariates, including TAC, systemic vascular resistance, aortic root dimensions, RV function, and pulmonary and aortic regurgitation. Overall, the fixed effects explained 35% of total LVEF variance (Nakagawa marginal R²=0.35), rising to 72% when patient-level random effects were included (conditional R²=0.72). The between-patient variance remaining after adjustment indicates that unmeasured stable patient characteristics, such as intrinsic myocardial factors, contribute substantially to differences in LVEF across patients.

*Sensitivity Analyses*

Study era (pre-2010, 2010–2014, 2015–2019, 2020 onwards) was not independently associated with LVEF (p≥0.19 for all era terms). Pulmonary valve intervention was not associated with LVEF (β=−0.41, p=0.48). Although PVI reduced pulmonary regurgitant fraction from 41% to 12% in patients with pre- and post-intervention imaging (p<0.001), this haemodynamic improvement did not translate into a detectable change in LVEF. All primary findings were unchanged after adjustment for either era or intervention status.

After adjustment for body surface area to account for growth over time, no systematic temporal drift was observed in systolic blood pressure, diastolic blood pressure, pulse pressure, or stroke volume. Mean age at study increased from 12.5 ± 5.2 years in 2003–2004 to 27.7 ± 13.1 years in 2025–2026, reflecting the maturation of the cohort and a shift toward a higher proportion of adult imaging.

A gradual increase in the ratio of sinus diameter to sinotubular junction diameter was observed over the study period (0.006 per year, p<0.001), consistent with a known shift in sinus measurement convention from cusp-commissure to commissure-commissure. STJ and ascending aortic dimensions showed no temporal drift. After calibration of sinus dimensions using the sinus-to-STJ ratio, the within-patient sinus coefficient was modestly strengthened (β=−0.29, p<0.001), while all other coefficients were unchanged.

1.	The fourth report on the diagnosis, evaluation, and treatment of high blood pressure in children and adolescents. *Pediatrics*. Aug 2004;114(2 Suppl 4th Report):555-76.

2.	Flynn JT, Kaelber DC, Baker-Smith CM, et al. Clinical Practice Guideline for Screening and Management of High Blood Pressure in Children and Adolescents. *Pediatrics*. 2017;140(3)doi:10.1542/peds.2017-1904

**Supplemental Tables**

**Supplemental Table 1. Variance Decomposition of Time-Varying Covariates**

| **Variable** | **N obs** | **Overall SD** | **Between SD** | **Within SD** | **ICC** |
| --- | --- | --- | --- | --- | --- |
| Compliance, mL/mmHg | 1,106 | 0.54 | 0.28 | 0.46 | 0.27 |
| SVR, mmHg·min/L | 1,101 | 5.58 | 3.80 | 4.09 | 0.46 |
| Sinus diameter, mm | 1,312 | 5.88 | 4.99 | 3.44 | 0.68 |
| RV ejection fraction, % | 1,447 | 8.45 | 7.06 | 4.89 | 0.68 |
| Pulmonary regurgitant fraction, % | 1,407 | 15.56 | 11.50 | 10.39 | 0.55 |
| Aortic regurgitant fraction, % | 1,408 | 5.38 | 4.13 | 3.61 | 0.57 |

Between SD = √var(random intercept); Within SD = √var(residual) from variance-components models with patient as random effect. ICC indicates intraclass correlation coefficient; MPA, main pulmonary artery; RV, right ventricular; SVR, systemic vascular resistance.
